# Spatial clustering in vaccination hesitancy: the role of social influence and social selection

**DOI:** 10.1101/2022.01.11.22269032

**Authors:** Lucila G Alvarez-Zuzek, Casey Zipfel, Shweta Bansal

## Abstract

The phenomenon of vaccine hesitancy behavior has gained ground over the last three decades, jeopardizing the maintenance of herd immunity. This behavior tends to cluster spatially, creating pockets of unprotected sub-populations that can be hotspots for outbreak emergence. What remains less understood are the social mechanisms that can give rise to spatial clustering in vaccination behavior, particularly at the landscape scale. We focus on the presence of spatial clustering, and aim to mechanistically understand how different social processes can give rise to this phenomenon. In particular, we propose two hypotheses to explain the presence of spatial clustering: (i) *social selection*, in which vaccine-hesitant individuals share socio-demographic traits, and clustering of these traits generates spatial clustering in vaccine hesitancy; and (ii) *social influence*, in which hesitant behavior is contagious and spreads through neighboring societies, leading to hesitant clusters. Adopting a theoretical spatial network approach, we explore the role of these two processes in generating patterns of spatial clustering in vaccination behaviors under a range of spatial structures. We find that both processes are independently capable of generating spatial clustering, and the more spatially structured the social dynamics in a society are, the higher spatial clustering in vaccine-hesitant behavior it realizes. Together, we demonstrate that these processes result in unique spatial configurations of hesitant clusters, and we validate our models on both processes with fine-grain empirical data on vaccine hesitancy, social determinants, and social connectivity in the US. Finally, we propose, and evaluate the effectiveness of, two novel intervention strategies to diminish hesitant behavior. Our generative modeling approach informed by unique empirical data provides insights on the role of complex social processes in driving spatial heterogeneity in vaccine hesitancy.

## Introduction

Vaccination is the single most effective way of mitigating the consequences of infectious diseases, and has resulted in steep declines in infection morbidity and mortality for vaccine-preventable diseases (VPDs) [1, 2]. To manage VPDs, high levels of vaccination coverage are critical to controlling outbreaks. However, vaccine uptake and the maintenance of herd immunity have been threatened in recent decades due to vaccine hesitancy behavior, which is the delay or refusal of vaccination despite its availability, and is influenced by factors such as complacency, convenience and confidence [3]. Vaccine hesitancy behavior has increased worldwide, leading the WHO to declare it as one of the top global health issues [4]. In high-resource settings like the United States, vaccine hesitancy for childhood vaccinations has been found to be concentrated in major metropolitan areas with substantial evidence linking recent VPD outbreaks of measles and pertussis in urban populations with hesitant behavior [6, 7], and more recently, has been jeopardizing the suppression of the COVID-19 pandemic [8, 9, 10, 11]. Continued work is thus urgently needed to better understand the characteristics and dynamics of hesitancy behavior so we may stem the tide of increasing hesitancy and prevent the threat it poses.

A critical complexity to our ability to diminish growing hesitancy is the distribution of the behavior within a population. Vaccine hesitancy has been shown to not be randomly distributed in populations but instead occurs in clusters of individuals in the same geographic areas. This geographic heterogeneity poses a major challenge to the control and elimination of VPDs, as it leaves pockets of unprotected sub-populations that may be vulnerable to outbreaks, even if they are part of a population with high average vaccination coverage. Past research has identified spatial clustering of vaccine hesitancy and geographic overlap between hesitancy clusters and clusters of reported disease cases [12, 13, 14]. Modeling work has also demonstrated that the presence of geographic clustering in hesitancy can drive increased disease emergence risk and transmission [15, 16, 17]. Such spatial heterogeneity in hesitancy can be driven by complex social processes but our understanding of the structure and impact of such processes remains limited.

Past meta-analytic work has proposed two broad categories of social determinants of hesitancy behavior in individuals (distinct from vaccine-specific issues such as safety or cost): contextual influences, arising from social, cultural, environmental and geographic factors; and group influences, arising from personal perception or the context of a social or peer environment or experience [3]. Based on these determinants, two social processes have been proposed that may generate behaviors such as vaccine hesitancy: a) *social selection*: in which socio-cultural determinants independently drive hesitancy behavior. So individuals that share those social characteristics interact with each other due to shared traits and independently engage in hesitancy behavior [18, 19, 20]; and b) *social influence*: in which vaccine hesitancy behavior propagates over existing social networks. So individuals that engage in hesitancy behavior influence others among their social contacts to do the same [21, 22].

Significant work has considered social selection and social influence theoretically and empirically for vaccine hesitancy [23, 24, 25] as well as other behaviors [26, 27, 28, 29]. However, all previous studies have focused on the individual-scale, in which individual attributes or behavior diffusion among individuals drives changes in behavior. What remains to be considered are the dynamics of social selection and social influence at the landscape-scale (i.e. across a large spatial area like an entire country made up of individual communities), in which the attributes of communities and collective behavior diffusion due to social connectivity between communities might drive dynamics of hesitancy behavior. As public health strategies are driven by a population health perspective, and because the geographic distribution of vaccination is critical to disease emergence across spatial scales, it is important to study dynamics at the landscape-scale beyond individuals [14]. At the same time, given the importance of spatial heterogeneity, considering vaccination at a fine spatial scale is also critical as vaccine uptake can vary widely within and between large geographical areas and large-scale aggregation of vaccination can mask local vulnerabilities [30, 16, 31].

In this study, we aim to characterize the role of social selection and social influence leading to spatial clustering in vaccination hesitancy at the landscape scale. We hypothesize that landscape-level spatial clustering in hesitancy may be produced by (a) social selection: geographically proximate communities may independently adopt similar behaviors because they share characteristics that promote the behavior; or (b) social influence: diffusion of vaccination behavior between socially proximate areas through learning of social norms and practices. In the case of social influence we expect to see that clustering in vaccination behavior arise surrounding highly hesitant populations, whereas under social selection, the clustering in vaccination behavior only reflects geographic clustering in underlying drivers. Using a fine-grain spatial network framework of the entire United States in which we consider the social and physical interactions between communities, we use a generative approach to introduce a specified level of social selection (based on community socioeconomic characteristics) or social influence (driven by the direct diffusion of behavior). Based on a distribution of hesitancy behavior, we then classify communities according to vulnerability to disease emergence: communities with a level of vaccine hesitancy putting them below the vaccination herd immunity threshold are considered vulnerable otherwise, they are considered protected. For a given proportion of vulnerable nodes, we mechanistically seek to understand whether social influence or social selection can independently (and together) generate spatial clustering patterns, and to what degree. We additionally seek to understand how the structure of the network over which these social processes occur impacts the generation of spatial clustering in hesitancy. We empirically validate our models using fine-grain data on vaccine hesitancy, social determinants, and social connectivity in the US. And, based on our findings, we propose and evaluate strategies to diminish overall levels of hesitancy in communities, as well as reduce spatial clustering in hesitancy. Our work integrates theoretical models with empirical data to develop a mechanistic understanding of the dynamic processes that potentially generate geographic clustering in hesitancy behaviors and informs interventions to ameliorate this dangerous behavior.

## Methods

Our work aims to characterize how social processes affect the spatial distribution of vaccine hesitancy at a landscape-scale and translate this understanding for mitigation strategies to diminish the spatial clustering present in hesitancy behavior across the US. In order to achieve this, we develop a theoretical spatial network model that allows for tunable spatial structure and social process dynamics. We parameterize this model with empirical data to validate our findings, and use a simulation approach to assess the effectiveness of proposed mitigation strategies.

### Empirical data

We integrate four empirical data sources into our work for model parameterization, validation and application, further described below: (1) we specify an infection case study, and use relevant empirical vaccine hesitancy data based on school exemptions; (2) from the US Census, we identify each county’s socioeconomic attributes relevant to our social selection model; and (3) based on social media activity data, we define social connectivity between communities relevant to our model of social influence. All at a granular landscape scale.

#### Vaccine hesitancy data

The principles of herd immunity suggests that if a large portion of a community is immune to disease due to vaccination, further transmission is unlikely. The proportion of the population that needs to be vaccinated to achieve herd immunity depends on how contagious the pathogen is. For our work, we focus on measles as a case study. Because measles is highly transmissible (*R*_0_ *≈* 20), the herd immunity threshold, *ρ*, for measles is *ρ* = 1 − 1*/R*_0_ = 0.95 [32]. Thus, an estimated 95% of a community needs to be vaccinated to achieve herd immunity for measles (we refer to such communities as *protected*). On the other hand, any community that cannot achieve 95% vaccination coverage due to vaccine hesitancy can expect sustained outbreak, we refer to these communities as *vulnerable*. State-mandated school entry immunization requirements in the United States play an important role in achieving high vaccine coverage for measles, but variations in vaccine exemption policies result in a patchwork of vaccine coverage across the country. All states allow exemptions for medical reasons and the vast majority also allow parents to opt out of childhood vaccination mandates due to non-medical religious or philosophical reasons. To measure vaccine hesitancy, we thus use data on non-medical exemptions for school children (Figure S2) [33], which is available for download at [34]. These data were collected from state health departments, and have been collated into a unified dataset at the county-yearly level for 24 states in the US. Thus, each county in the US is characterized by a level of hesitancy, strongly correlated with vaccine coverage.

#### Socioeconomic data

Previous work has shown that hesitancy behavior for childhood vaccinations such as the measles-mumps-rubella (MMR) vaccine is associated with socioeconomic traits. In particular, smaller average household size and larger average household income levels have been shown to be positive correlated with to increased hesitancy behavior [18, 35, 36]. We thus use these two socioeconomic traits in our measles case study, based on county-level data from the US Census (shown in Figure S5).

#### Social connectivity data

To model empirical social connectivity between communities (particularly for the social influence process), we use the Facebook social connectedness dataset [37], which is available for download at [38]. A pair of communities (i.e. US counties) are connected if Facebook users in one community are “Facebook friends” with users in the other community. The social connectedness index (SCI) captures the strength of the interaction. We created SCI-thresholded networks (SCI *>* 400) based on a weighted network analysis S1. At SCI *>* 400, we find that the average degree of 6 and contains 2496 counties.

We also find that the structure of the social connectivity is both spatial (i.e. communities are connected with their geographic neighbors) and aspatial (i.e. communities are connected with geographically distant communities). While social media connectivity does not capture all social connections between communities, the significant spatial structure of this network suggests that this connectivity may reflect interactions between communities in the physical world which tend to be geographically structured. The structure of this data also suggests that the landscape social network may be well approximated by a small-word network model, which represents networks with significant spatial connections and occasional aspatial connections. To confirm this, we measure the small-worldness coefficient *σ ≈* 178 (≫1) and ensure the empirical network exhibits small-world characteristics [39, 40]. We will thus use a Watts-Strogatz small-world network as the structure of our theoretical model. For most of our analyses, we will evaluate the entire range of small-world structures, from fully spatial (rewiring probability, *p* = 0) to fully aspatial (*p* = 1). However, for parameterization, we also infer the rewiring probability of the social connectivity data. To do so, we estimate the average clustering coefficient (*c* ≃ 0.29) and the average shortest path length (*l* ≃ 3.8) of the social connectivity network at a threshold of *SCI* > 400, and compare it to the the network properties of a small-world network with varying rewiring probabilities. From this, we estimate a small-world rewiring probability of *p* ≈ 0.2 for the social connectivity dataset.

#### Empirical estimates for model parameterization and validation

We aggregate data from 2015 and 2018 for vaccine hesitancy, socio-economic traits and social connectedness (as described above). Because the datasets have non-overlapping coverage, we arrive at data on 184 counties (nodes), belonging to a total of 4 states (Arizona, California, Maine, and Virginia). (We note that these counties are a convenience sample based on data availability, and are not necessarily representative of the entire country.)

From this integrated dataset, we estimate parameters necessary for model parameterization. We estimate that vaccine hesitancy behavior at the county level follows an exponential distribution, *P*_(*η*)_ = *λ e^−λη^* (Figure S4). We also find that hesitancy behavior has been increasing nationally over recent years: 9% ± 0.2% of counties in our dataset were vulnerable (i.e. the proportion of the population not hesitant is below the herd immunity threshold) in 2015, which increases to 25% ± 1% of counties as vulnerable in 2018. At the community level, we estimate an average increase in hesitancy per county of 1% from 2015 to 2018 (Table S1).

For model validation, we estimate parameters for social selection and social influence, and simulate the social influence process on the empirical dataset. We estimate the empirical social selection parameter (*β*\*) based on the empirical distribution of socio-economic traits (average household size and average household income) and the definition of the parameter as specified below. We estimate the empirical social influence parameter (*α*\*) as the average hesitancy level of the social contacts (as defined by the social connectivity data) of nodes which were observed to increase in hesitancy from 2015 to 2018. For both parameters, we produce uncertainty estimates using 1000 bootstrap networks of 70% of the original network size (184 counties). We also calculate the empirical spatial clustering as defined below using 1000 bootstrap samples of the original sample size (184 counties).

### Generative landscape network model

We consider a society made up of spatially and socially interconnected communities. We represent this society as a landscape-level network in which communities (i.e. US counties) are represented by network nodes and spatial proximity or social interactions between communities are represented by network edges (Figure 1a). We define spatial proximity based on shared land borders between US counties.

**Figure 1:**
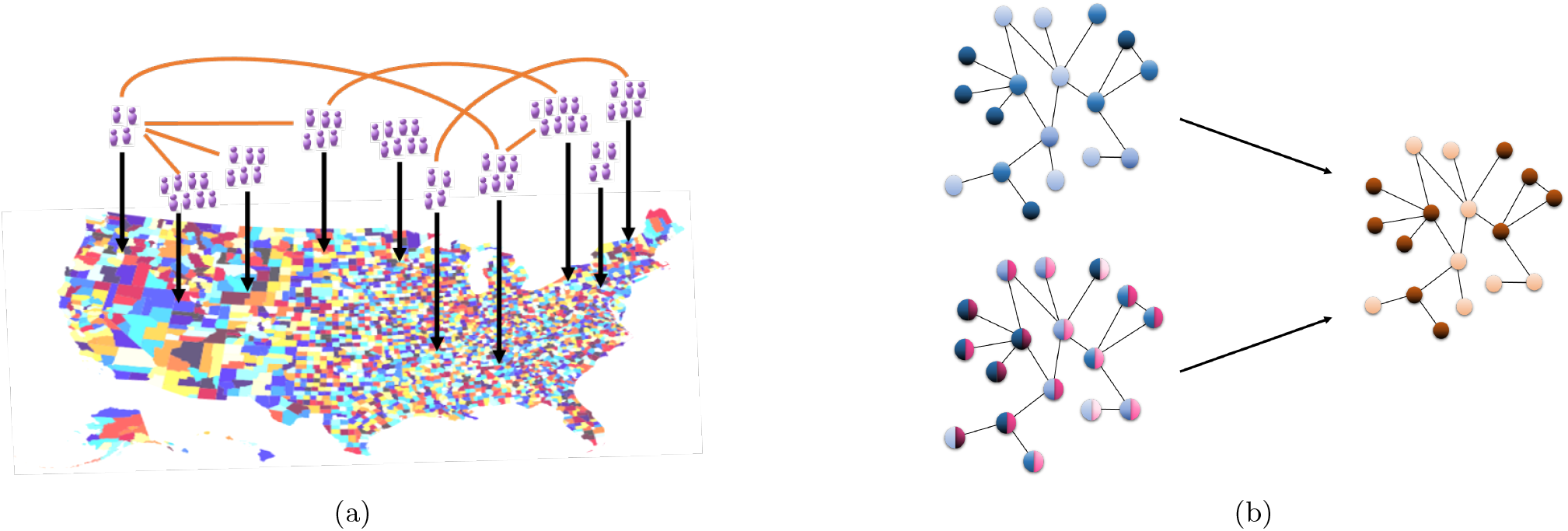
(a) A schematic of the landscape-level spatial network in the United States. Counties, which contain populations of individuals, are represented by nodes, and interactions between the county populations determine the edges of the network. (b) A schematic of the change in vulnerability due to the social influence and social selection processes. Under social influence (top arrow), each community (node) starts with a level of hesitancy (denoted in blue in the top network), and hesitancy behavior spreads through the network across social interactions (edges) between communities from hesitant (dark blue) communities to non-hesitant ones (light blue). Under social selection (bottom arrow), communities with similar traits (denoted in pink in the lower left network) tend to be connected, and some traits independently lead to hesitancy behavior (denoted in blue in the bottom left network). Each process can give rise to the same distribution of vulnerability to outbreaks (dark orange = vulnerable community, light orange = protected community in the right network).

We develop our landscape network model to capture structure relevant to the two social processes of interest. Social selection at the landscape-level describes the tendency of communities with similar attributes being geographically proximal and connected. As individual and community attributes have been demonstrated to be associated with vaccine hesitancy behavior, social selection leads to spatial clustering in hesitancy due to spatial clustering in attributes. Conversely, social influence at the landscape-level describes a diffusion of hesitancy behavior over social connections between communities. These social links can be spatial (due to physical mobility, e.g. commuting behavior) or aspatial (due to social interactions, e.g. online social media) in nature. Such network structure is theoretically well-represented by the Watts-Strogatz small-world network model [41], which allows for tunable spatial network structure while preserving other network structure features such as average connectivity. For our theoretical characterization of the role of social selection or social influence in generating spatial clustering, we generate an ensemble of Watts-Strogatz small world networks of size *N* = 2048, average degree *k* = 6 (informed by the analysis of the empirical social connectivity data), and spatial structure varying from spatial (*p* = 0) to aspatial (*p* = 1) to evaluate a range of network structures.

For each social process, we use a Monte Carlo approach to generate varying levels of the process on our spatial network model. Then we consider how the distribution of hesitancy due to social selection versus social influence leads to spatial clustering, or pockets of high hesitancy communities in geographic proximity. For social selection, we keep the proportion of vulnerable nodes constant, but shift the configuration of vulnerability, driven by homophily in attributes. For social influence, hesitancy behavior is propagated between communities, directly increasing the proportion of vulnerable communities. In order for the two processes to be comparable, we restrict the proportion of vulnerable communities that can be generated through social influence to match the proportion assumed for social selection.

To measure spatial clustering, we calculate the level of spatial assortative mixing [42] in the community herd immunity status (vulnerable/protective) where the network is the landscape-level spatial network (i.e. edges between communities due to spatial proximity). Hence, we use

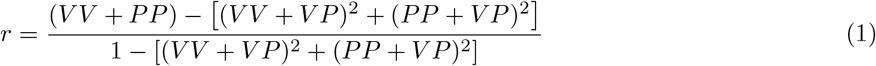

where *V V* measures the fraction of all spatial proximity edges that are between vulnerable nodes, *PP* is the fraction of all spatial edges that are between protected nodes, and *V P* measures the fraction of all spatial edges that are between vulnerable and protected nodes.

Thus, high levels of spatial clustering indicate a strong tendency of vulnerable (protected) nodes to be connected creating pools of susceptible individuals.

### Modeling social selection at the landscape scale

We model social selection in a landscape-scale spatial network in which communities are connected to each other due to geographic proximity, and community attributes are modeled based on a level of social selection. In particular, each network node (community) is described by a set of attributes associated with hesitancy behavior (representing, for example, average household income and average household size).

We begin by initializing each node with a level of vaccine hesitancy, based on an exponential distribution (*η* ∼ Exp(*λ*)) based on the observed data. We parameterize lambda using the observed distribution of hesitancy in 2018 (with approximately 25% of all communities as vulnerable). The vector of attributes for each node is then defined by ***X*** = *η*^1*/γ*^. This makes the traits Weibull distributed with the following parameterization: ***X*** ∼ Weibull(***γ***, *λ*^−1^) (more details are in the Supplement).

We then measure changes in the attribute distribution across nodes using the Mahalanobis distance, *µ*, which is defined as a dissimilarity measure between two random vectors of the same distribution with a covariance matrix ***S***. For a network with a set of edges, *E*, and attributes ***X***_*i*_ for node *i*, we defined the global distance in the network as:

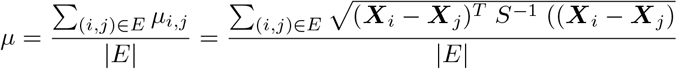

Based on this distance, we define the *social selection* parameter, *β*, as the deviation in distance in the simulated scenario (*µ*) from the distance in a random scenario 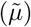 in which attributes are distributed randomly and independently, without correlation: 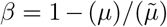. Our model aims to reach a desired level of social selection, *β*\* and achieves this through an attribute randomization algorithm. The attribute vectors of nodes are swapped, and if the swap succeeds in increasing *β*, it is retained. Attribute randomization continues till *β* = *β*\*. Additional model details can be found in the Supplementary Methods.

### Modeling social influence at the landscape scale

We propose a social influence model that simulates diffusion of hesitancy behavior via social contacts between communities. Our model is inspired by bootstrap percolation [43], and it proceeds as follows: Each community (network node) is initialized with a level of hesitancy *η* sampled from an exponential distribution. Nodes are classified based on the herd immunity threshold (*ρ*) as vulnerable (*η* ≥ (1 − *ρ*)) or protected (*η* < (1 − *ρ*)). For our simulations, we choose measles as a case study and assume *ρ* = 0.95. At each time step, each node *i* has it’s own level of hesitancy and is exposed to the hesitancy levels of its neighbors, *j*. If the average hesitancy level for the node and its neighbors (avg({*η*_*i*_, *η*_*j*_ ∀*j*})) exceeds a tolerance (*α*), then the node’s hesitancy increases by *δ* (which we parameterize as 0.01 based on the observed hesitancy data). We refer to *α* as the *social influence parameter*, and define it as the minimum exposure required to increase a community’s hesitancy. For any protected nodes, if their new hesitancy level exceeds 1 − *ρ*, then they become vulnerable.

Our model is initialized with 10% of all communities being vulnerable and hesitancy propagation is permitted till 25% of all communities are vulnerable (parameterized based on our observed hesitancy data). This restriction is added so as to make the social influence process results comparable to those from the social selection process.

### Mitigation Strategies

Understanding the role of social selection and influence in the spatial clustering in vaccine hesitancy allows us to develop effective intervention strategies to reduce both hesitancy and spatial clustering in hesitancy. We design strategies where specific communities can be targeted to reduce hesitancy levels. We evaluate this strategy on a theoretical social network with spatial structure parameterized based on the Facebook social connectedness dataset as a Watts-Strogatz small-world network with rewiring probability *p* = 0.2 (as described in the Empirical data section). We measure the effectiveness of each strategy in terms of a reduction in spatial clustering compared to the control case of no action.

#### Reducing clustering through social selection

For our first strategy, we propose to reduce clustering caused by social selection. Because social selection relies on a significant association between community attributes and hesitancy, we seek to target the distribution of attributes themselves. In particular, our strategy aims to select a potential vulnerable county given its level of attributes (which is positively correlated with hesitancy) and the conformity in that attributes among neighboring communities. Thus, for each county *i* we define an index, *ζ*_*i*_, as,

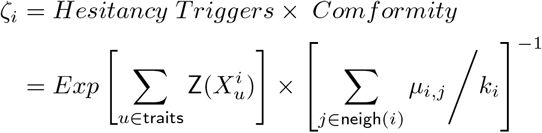

where 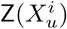 is the z-score of the attribute *X*_*u*_ for the focal county *i*, and *µ*_*i,j*_ is the Mahalanobis distance between the attributes *i* and each neighbor and measures the conformity of the neighborhood of *i*. To implement the strategy, we target nodes with the 10% highest *ζ* values. Note that target implies changing vulnerable status for protection. In absence of hesitant data, the primary purpose of this strategy is to efficiently detect vulnerable counties to invest resources to break hesitant clusters.

#### Reducing clustering through social influence

For our second strategy, we focus on our results which have demonstrated that small changes in network structure have a substantial impact on spatial clustering in hesitancy caused by social influence. Thus for this strategy, we aim to reduce clustering caused by social influence by altering the network structure: specifically, we target and reroute edges (i.e. social interactions over which social influence occurs) between vulnerable and protected communities with probability *ω* (constant across all edges). The rerouting strategy is carried out within the social influence model, and occurs whenever a node changes status to becoming vulnerable (due the ongoing influence process). Any edges that the vulnerable node has with protected nodes are disconnected from the vulnerable node and rerouted to a randomly selected protected node (so a vulnerable-protected edge is replaced with a protected-protected edge). In this way, further propagation of hesitancy from the vulnerable node to protected nodes can be minimized.

## Results

### Spatial clustering can be generated by social selection or social influence

Social selection and social influence have been demonstrated to be possible mechanisms driving vaccination hesitancy among individuals. Here, we examine theoretically how these two social processes can generate spatial clustering at the landscape scale. To achieve this, we consider Watts-Strogatz small-world networks with tunable spatial structure, and develop two mechanistic models to generate a tunable level of social selection or social influence in these networks. With this generative approach, we seek to characterize spatial clustering under the two social processes and in light of different degrees of spatial structure.

In Figure 2, we illustrate that the spatial clustering can be generated by either social influence or social selection. For both processes, low values of influence/selection result in little spatial clustering in vaccination hesitancy; but as the intensity of either process increases, spatial clustering also increases. Additionally, in both cases, increased spatial structure tends to favor spatial clustering, and small changes in the structure of community connectivity lead to proportional changes in clustering with social influence, while moderate to high levels of spatial structure do not drastically change spatial clustering with social selection.

**Figure 2:**
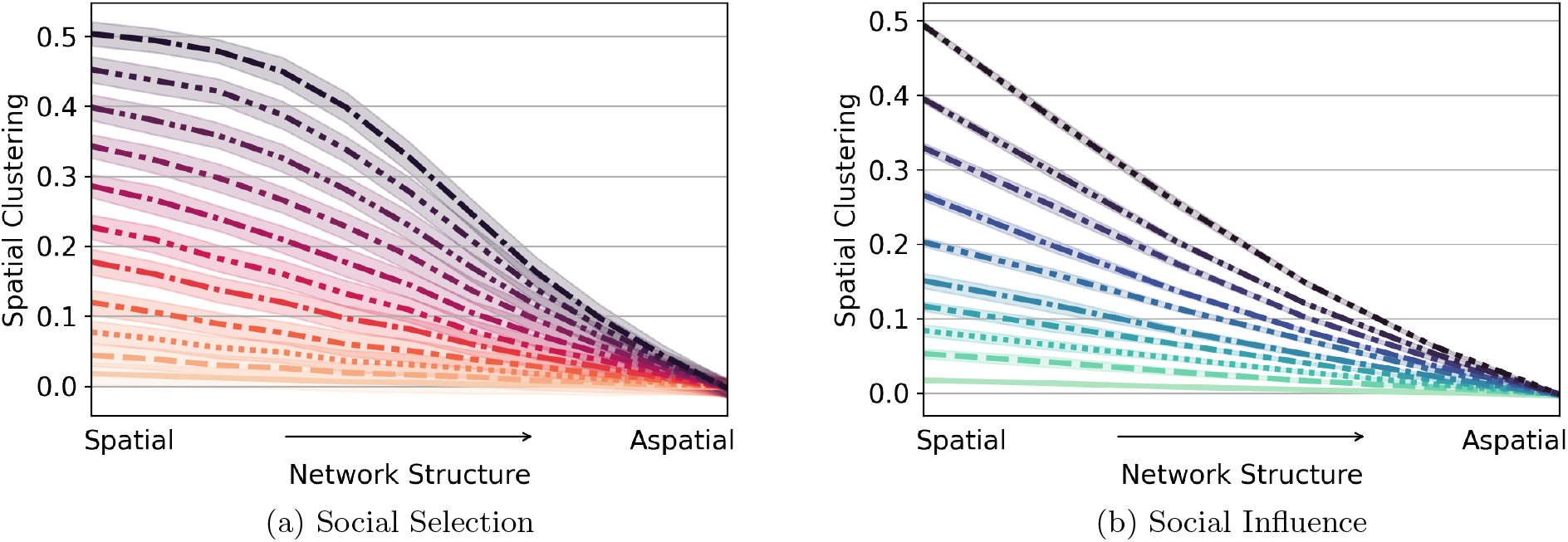
Spatial clustering generated by social selection (left) and social influence (right). For each processes we show the spatial clustering in vaccine hesitancy as a function of the network structures, from highly spatial (ring) to aspatial (random) networks. Each curve corresponds to a different intensity of the given social process, from high (top, darker shades) to low (bottom, lighter shades).

We also analyzed how the spatial nature of networks influenced maximum outbreak size (Figure S8). Specifically, we evaluated how the spatial structure in the network impacts the reach of a possible disease outbreak, due to the spatial clustering of hesitant behavior. Even though both social processes have different mechanisms generating their spatial clustering, we found that the maximum possible outbreak size is similar, although there is much more variation for the case of social influence.

### Social selection increases spatial clustering only in affectable societies

Social selection and social influence are driven by different social mechanisms and are expected to co-occur in complex societies. Thus, we next develop a model to consider the presence of social selection and social influence simultaneously to generate spatially clustered hesitancy behavior. Our model starts with a desired level of social selection, and allows hesitancy behavior to spread using our social influence model. With this model, we explore two types of societies: i) an affectable society is one with a low social influence parameter (*α*), in which individuals are receptive to anti-vaccine messaging and quickly adopt hesitant behavior, and ii) a determined society with a high social influence parameter, in which individuals are resistant to anti-vaccine messaging, and are less likely to adopt hesitant behavior. In all cases, we allow the influence process to continue until a maximum (25%) of communities are vulnerable as we are interested in understanding the spatial configuration of the same proportion of vulnerable communities under different scenarios.

In Figure 3, we show the level of spatial clustering generated by social selection combined with social influence in the affectable versus determined societies. In an affectable society (low social influence parameter *α*), individuals tend to adopt hesitancy behavior easily, and we find that social selection has a strong impact in determining spatial clustering, particularly for more spatially structured societies. For a society with high social selection, as clusters of high hesitancy already exist, social influence favors their increase. For low values of social selection in an affectable society, there are few clusters, and as the population tends to turn hesitant easily (low social influence parameter), numerous small clusters are rapidly created throughout the network. In a determined society (high social influence parameter *α*), on the other hand, communities are more resistant to adopt a behavior against vaccines; to become hesitant, high consensus is needed for hesitancy among a community’s neighbors. Therefore, only few larger cluster sizes prevail.

**Figure 3:**
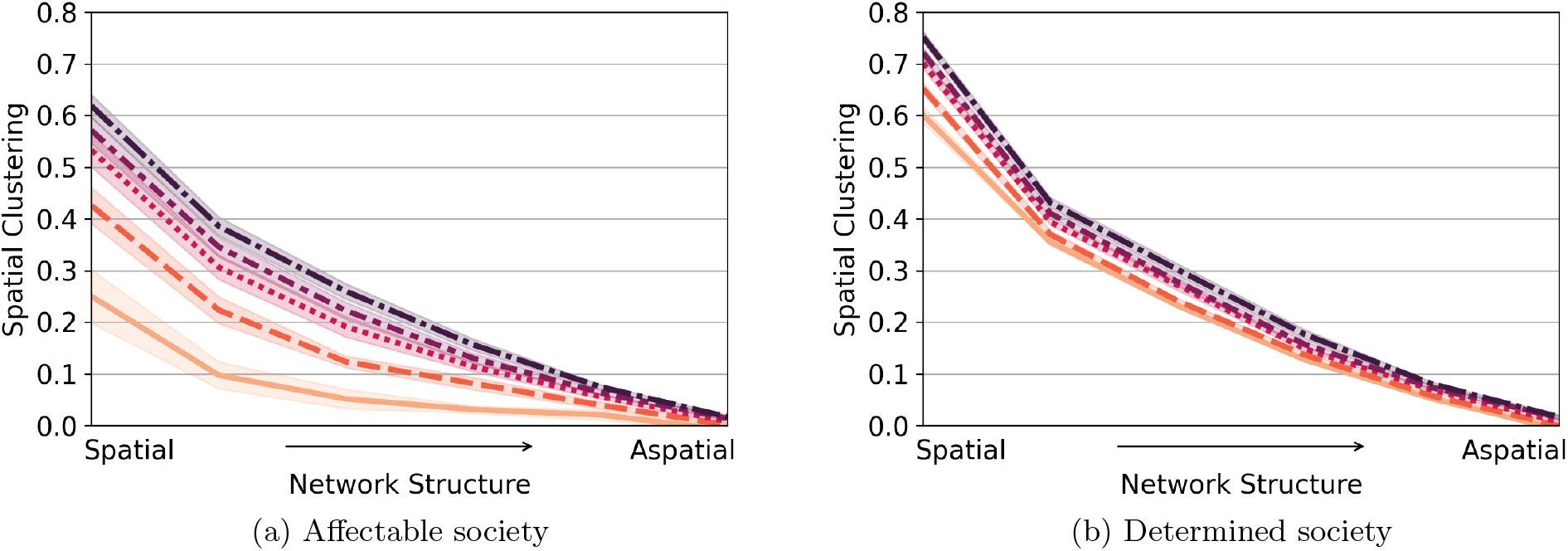
(a) An affectable society with *α* = 0.2, which has a low social influence parameter and (b) A determined society with *α* = 0.6 which as a high social influence parameter. Each curve corresponds to a different level of initial social selection; from homophilous (dark) to random (light) networks.

**Figure 4:**
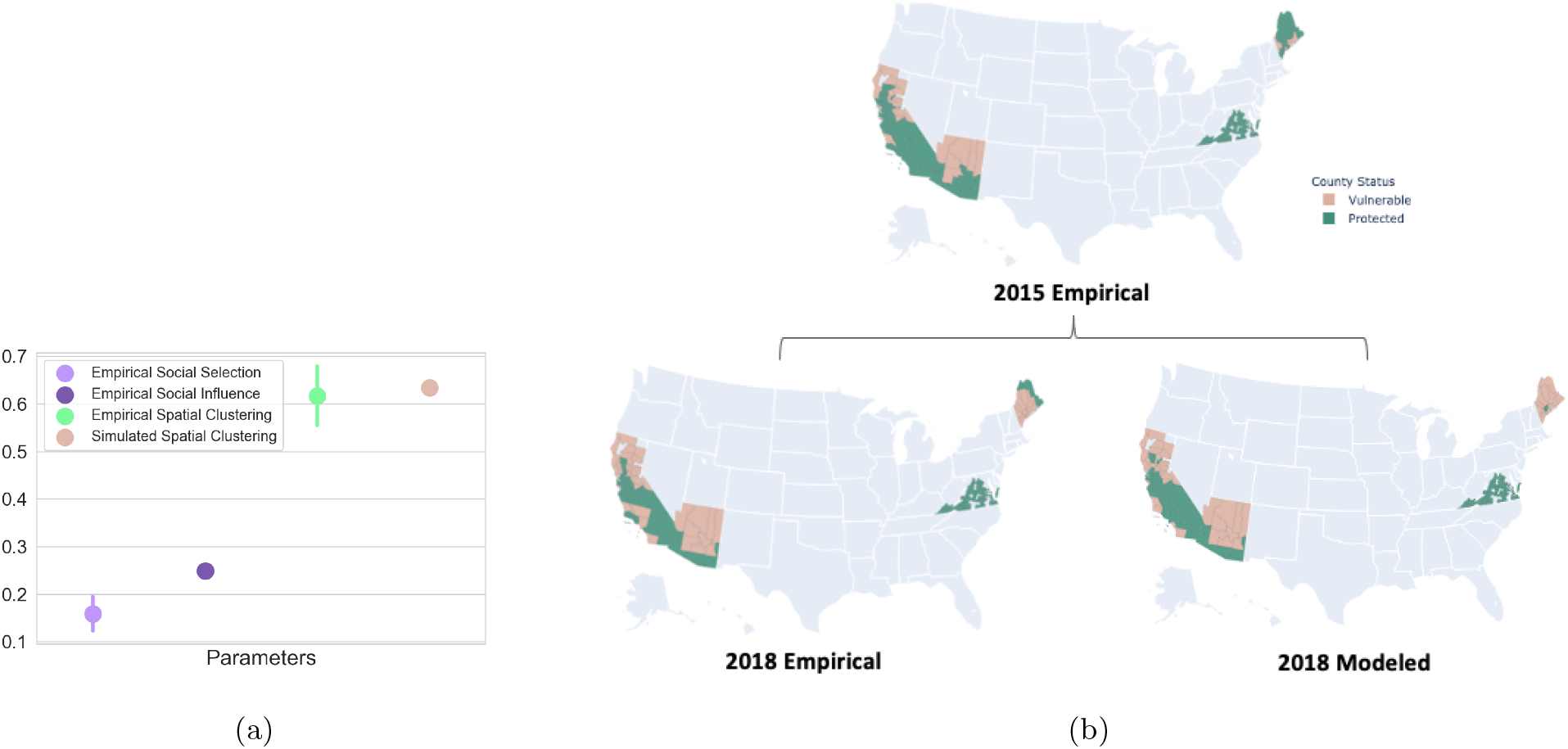
(a) We show estimated social selection *β*, social influence *α*, the spatial clustering values based on observed hesitancy data, and that from stochastic simulations of our theoretical model. b) Pink colored counties 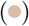 are vulnerable (high hesitancy) and green colored counties 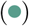 are protected (low hesitancy). We illustrate the observed data for 2015, alongside the observed and social influence modeled estimates for 2018.

**Figure 5:**
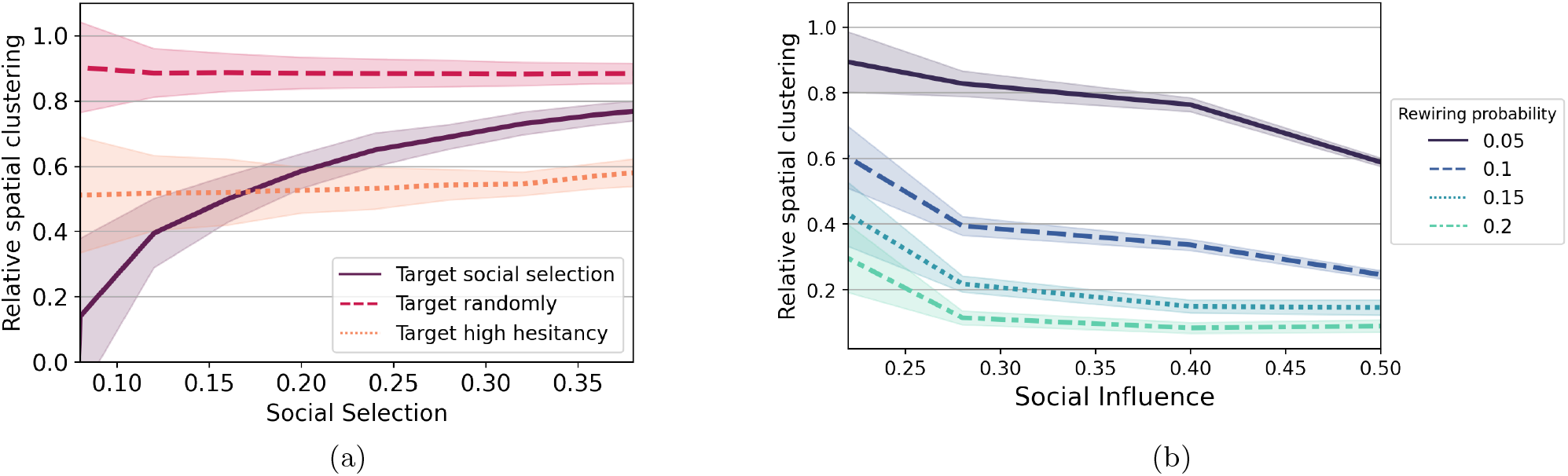
Reducing spatial clustering via interventions (a) social selection strategy: we target 10% of vulnerable counties in each strategy scenario; and (b) social influence strategy: each curve corresponds to a different probability of rerouting between a vulnerable/protected edge.

Furthermore, we find that in the affectable society, social selection plays an essential role in the dynamics, while in the determined society, the influence process is so strong that the initial configuration of the system is not significant. We can conclude that affectable societies are more prone to create a larger number of smaller cluster sizes. In contrast, skeptical societies result in a small number of larger cluster sizes and efficiently spread hesitancy.

### Our generative model captures observed patterns of hesitancy

We evaluate and validate our approach with observed vaccine hesitancy data based on childhood school exemption data from the states of California, Arizona, Maine, and Virginia during the years 2015 – 2018 [33]. To consider social selection, we characterize the empirical spatial distribution of the socio-economic traits of average income and household size shown to be associated with hesitancy behavior. To consider social influence, we use empirical data on social connectivity between US counties and the empirical distribution of hesitancy for childhood vaccination.

In Fig.4a, we estimate the social selection parameter *β* based on the geographic distribution of socioeconomic attributes to be *β*\* = 0.165. We also estimate the social influence parameter as *α*\* = 0.253. In the supplementary information, we highlight that empirically, influence is not homogeneous geographically but displays significant heterogeneity due to spatial clustering (Figure S8). We estimate the empirical spatial clustering for 2018 as 0.61 suggesting that the observed spatial distribution of hesitancy is heterogeneous.

To evaluate our social influence model, we consider the spatial extent and distribution of vulnerability as observed empirically and as estimated by our model. In Fig.(4b) we display the observed patterns of vulnerability in both 2015 and 2018. We compare these with the estimates of vulnerability from our social influence model based on a social influence parameter of *α* = 0.253. Compared statistically, the empirical and modeled (based on an average of 1000 model estimates) spatial distributions have an F-score of 0.925. We also estimate the modeled spatial clustering as 0.625, making it consistent with the observed spatial clustering. This comparison suggests that social influence is indeed capable of generating spatial configurations of vulnerability similar to observed spatial data, based on a complex social network which has both spatial and aspatial links. To highlight this point, Santa Barbara county in California (the southwestern most vulnerable county in 2018 in both the empirical and modeled maps) is shown to be not vulnerable nor spatially connected to vulnerable communities in 2015. However, in 2018, it becomes vulnerable due to social interactions with communities not in its geographic vicinity.

#### Spatial clustering in hesitancy can be diminished through interventions

Informed by an understanding of the impact of each social process on generating spatial clustering in hesitancy theoretically and empirically, we now propose and assess the effectiveness of intervention policies. The considered policies not only reduce the prevalence of hesitancy, but also reduce spatial clustering in hesitancy, with the goal of reducing pockets of vulnerable communities rather than simply eliminating isolated counties of high hesitancy.

Social selection is driven by two key features: (a) a strong correlation between a socioeconomic trait and hesitancy, leading to high levels of the trait driving high hesitancy; and (b) a conformity or similarity in traits among neighboring communities. Hence, we propose our first intervention strategy (‘target social selection’) to diminish spatial clustering in hesitancy behavior by targeting both high levels of hesitancy-correlated traits and conformity in those traits. Because vaccine hesitancy data is difficult to obtain and may not always be available, our proposed strategy offers an alternative without the need for intensive data.

To compare how effective our social selection strategy is, we compare it with: i) a best case scenario (‘target high hesitancy’) in which available hesitancy data is used to target communities with the highest levels of hesitancy; a worst case scenario (‘target randomly’) in which communities are targeted at random.

In Fig.5(a), we show that all strategies are effective in diminishing clustering relative to no intervention. Because the two comparison strategies do not act directly on spatial clustering, their impact does almost not change with increasing social selection. On the other hand, targeting social selection by targeting traits and conformity results in a larger reduction in spatial clustering for lower values of social selection. For high values of social selection, spatial clustering is higher with our ‘target social selection’ strategy than with the ‘target high hesitancy’ strategy. Within a context of high segregation, the best strategy is merely to target high hesitant counties, which implies having available hesitant data. If the level of social selection is relatively (as we find in our empirical analysis, *β* ≈ 0.16) our findings suggest that our strategy will perform similarly to targeting high hesitant counties.

For our second strategy, we exploit the impact of network structure on spatial clustering, and seek to reduce clustering by altering the structure of the social connectivity between communities, by rerouting social connections to make them less spatial. In Fig.5b(b), we find that as the social network is made less spatial, there is a larger reduction in spatial clustering, and that this impact increases with larger values of social influence.

## Discussion

Vaccination hesitancy is a dangerous behavior that threatens the maintenance of herd immunity, and spatial clustering of this behavior amplifies outbreak potential with when the behavior is rare. In this work, we have evaluated the role of social selection and social influence leading to spatial clustering in vaccination hesitancy. Previous literature have shown these two processes as strong candidates to be a mechanism behind this hazardous behavior. Thus far, these factors have been well studied at an individual level, and here we expand this individual-level focus to a landscape perspective, where influence and selection are processes occurring among communities within a country. Transitioning to this large population perspective is crucial as data collection is feasibly and routinely done at this scale, and state-level and national public health policies are designed and implemented at this scale. To achieve this, we use a complex network approach to describe the spatial structure of counties in the United States, and we develop two generative models that describe influence and selection. Our theoretical findings suggest that both social processes generate hesitant behavior clustering, but are configured differently. In network structures ranging from spatial to aspatial, social selection tends to be a more robust process where significant changes in the network structure are needed in order to impact the spatial clustering. On the other hand, spatial clustering in hesitancy driven by social influence depends exponentially on the network structure. Thus, small changes in the distribution of edges have an impact on the spatial clustering.

Past work has demonstrated that exposure to a hesitant neighborhood in online social networks leads to hesitant behavior, so we hypothesized that both social processes are likely to co-occur. Our theoretical findings of the combined processes suggest that when the a society trusts hesitancy propaganda, social selection plays an important role and many smaller clusters of hesitancy appear. On the other hand, when a society tends to be more skeptical about propaganda, social influence overcomes and a few larger clusters appear, despite the same overall frequency of vulnerable communities. Our empirical validation conversely highlights how social influence can take advantage of a society already affected by social selection to spread hesitancy easily, generating observed patterns of spatial cluster distribution. Understanding this difference in cluster configuration and distribution is critical to prediction of outbreak potential [16], and suggests the benefit of incorporating the social context of a community in public health programs.

To aid in the development of effective public health mitigation strategies, we take advantage of our theoretical results. We proposed two intervention strategies to reduce not just vaccine hesitancy levels but to reduce spatial clustering in hesitancy to have a disproportionate decline in outbreak potential. Our strategy to reduce clustering caused by social selection focuses on targeting communities that are vulnerable due to their own socio-economic traits but also are surrounded by a socio-cultural environment with a high tendency towards hesitancy; once the communities are identified, traditional public health measures of reducing vaccine hesitancy such as healthcare provider training and community health outreach programs can be implemented. Our results demonstrate that when social selection in a society is low, our strategy outperforms the strategy of directly targeting counties with known levels of high hesitancy because by simply targeting hesitancy, we may be reaching isolated highly- hesitant nodes surrounded by protected communities to whom they pose little danger. Additionally, we note that even in high-resource settings such as the US, fine-grain national data on vaccine hesitancy continues to be poorly measured and inaccessible [33], thus the advantage of our social selection strategy is that it does not require access to any direct data on hesitancy. On the other hand, the success of this strategy relies on studies that identify clear associations between community traits and hesitancy behavior. Significant such work exists based on survey and interview studies of parents, social experiments, and fine-grain ecological studies, and we must continue to invest in such work to characterize the evolving socio-cultural landscape of hesitancy. Our strategy to reduce clustering caused by social influence proposes to manipulate the spatial connectivity that underlines the social influence process for communities with observed high hesitancy. While such a strategy is likely to be impractical in traditional social networks, the social connectivity between communities due to social media usage may be amenable to manipulation by expanding upon the geo-targeted and connection- targeted digital marketing techniques that are already common. Such processes are also being considered for political mobilization [44] and health applications[45, 46], and future studies could experimentally evaluate the effectiveness of such a strategy [47].

Our work has some limitations. Our social influence model design is based on findings carried out in online social networks at an individual level. Even though there is some evidence of peer influence between communities for health behaviors, we believe more evidence in vaccination hesitant sentiment is needed. Due to the lack of data, we also assume that socio-economic traits remain constant, thus the level of social selection remains constant in our models. We do note that we expect social selection to be a slower process compared to social influence as socio-cultural environments tend to evolve slowly, so do not believe this to be an unrealistic assumption. Lastly, we recognize the need for more empirical validation of our findings, and we advocate for more data to be collected on vaccine hesitancy at a fine-grain and across the United States and for a range of vaccines. The COVID-19 pandemic has shone a new light on the benefits of vaccination and the dangers of vaccine misinformation, and we hope this leads to sustained attention on these important public health areas.

Understanding how social behavior impacts spatial clustering is challenging, but the methods and our findings are a step forward towards understanding the underlying processes that generate these clusters, both theoretically and empirically, and designing mitigation strategies to reduce clustering of vulnerable populations. Generative models of social behavior such as ours can also inform dynamical behavior-disease models which have been limited to assuming vaccine hesitancy in a non-spatial context and only through the lens of social influence [48]. The threat that vaccine hesitancy poses to local elimination of vaccine-preventable childhood diseases is growing, and we advocate for continued progress on mathematical modeling of this phenomenon from both a social and spatial perspective.

## Data Availability

All data produced in the present study are available upon reasonable request to the authors

## Acknowledgements

Research reported in this publication was supported by the National Institute of General Medical Sciences of the National Institutes of Health under award number R01GM123007. The content is solely the responsibility of the authors and does not necessarily represent the official views of the National Institutes of Health.

We also acknowledge feedback on this work from our collaborators Pej Rohani and Murali Haran.

## Supplementary Methods

### Empirical social network structure

To model empirical social connectivity between communities (particularly for the social influence process), we use the Facebook social connectedness dataset [37]. A pair of communities (i.e. US counties) are connected if Facebook users in one community are “Facebook friends” with users in the other community. The social connectedness index (SCI) captures the strength of the interaction.

**Figure S1:**
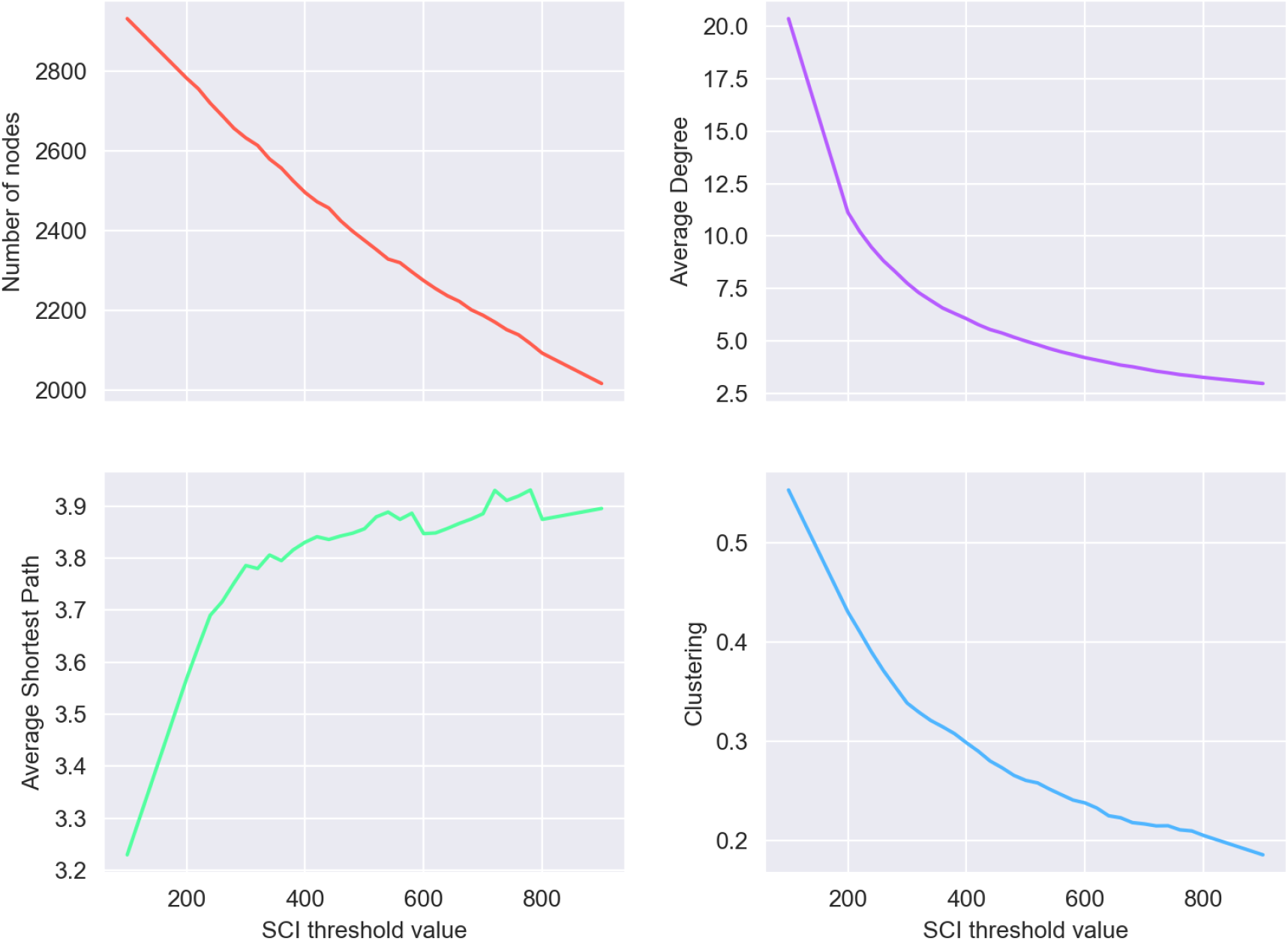
The Facebook network initially incorporated edges between every county in the US, including a vast number of very low weight edges. To only include consequential edges, we defined an edge weight threshold, excluding all edges that fall below this threshold value. To achieve this, we varied the edge weight threshold between 100 and 900, and, using the remaining network, calculated the included number of nodes (red), the average degree of nodes (purple), the average shortest path of the network (green), and the network clustering coefficient (blue).

The SCI has a heterogeneous distribution across the network and considering edges with all SCI values greater than zero results in an extremely dense network with little heterogeneity. Thus we use SCI-thresholded networks. To identify an SCI threshold, we consider the change in several network characteristics (average degree, average shortest path, and average clustering coefficient). We identify an SCI threshold of 400 as the smallest threshold above which network structure does not significantly change.

### Social Selection: simulation process

Our simulations code proceeds according to the following rules:

1. Randomly assign the vector of traits and hesitancy level to each node in the network
2. Calculate the level of social selection, *β*
3. Randomly choose two nodes in the network and swap their vector
4. Measure the new level of social selection, *β*\*
5. If *β*\* is closer to the desired value of social selection, keep swap. Otherwise, undo swap and go to step 3
6. If *β*\* is the desired value, stop. Otherwise, keep the swap and continue from step 3.

## Supplementary Figures

**Figure S2:**
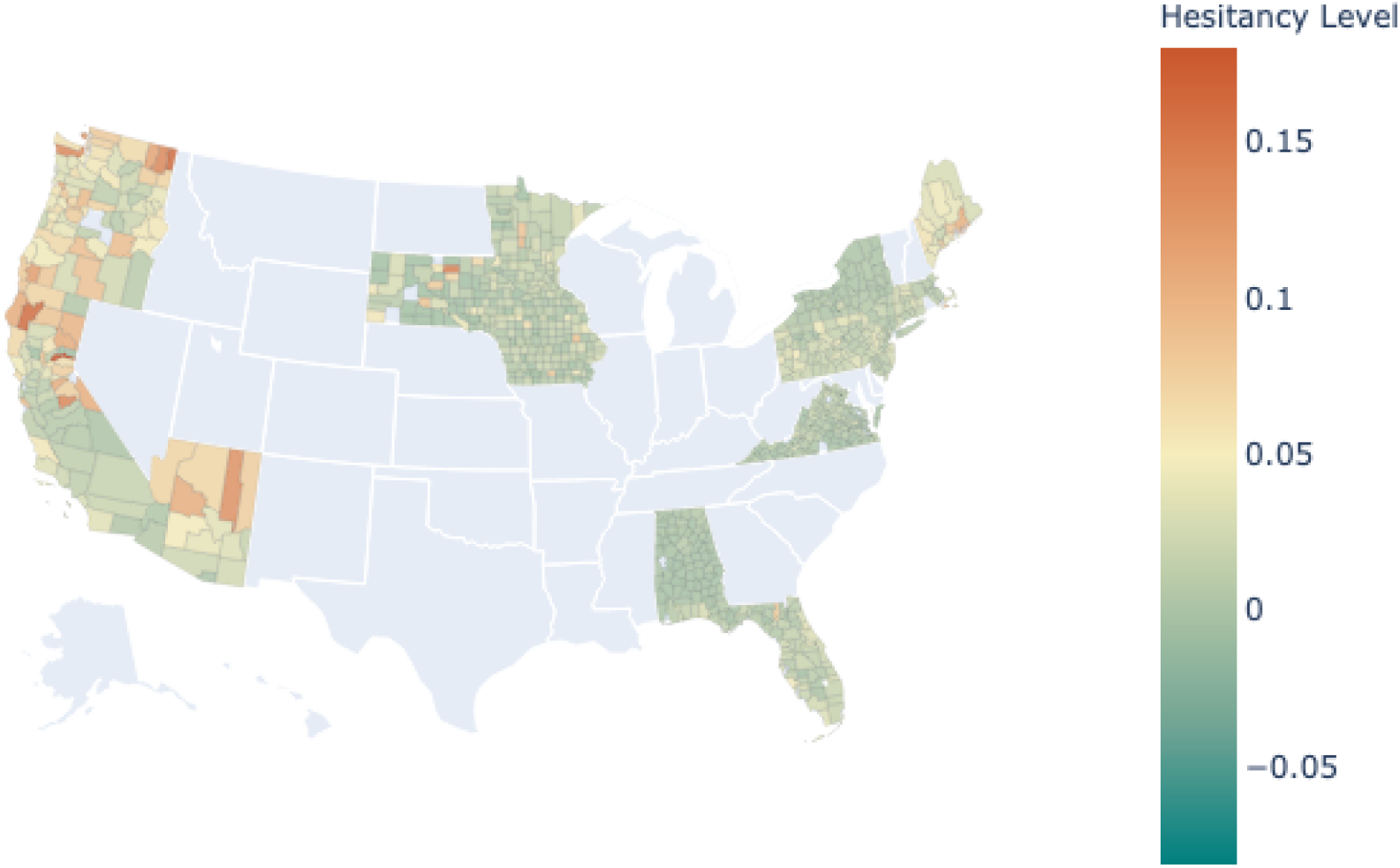
Map representation of the level of childhood vaccination hesitancy in the US in 2015 (proportion of kindergarten or 12th grade students with school vaccination exemptions). Note that counties in gray correspond to missing data

**Table S1:** Increment in hesitant behavior for the period of years 2015 – 2018.

|  | California | Virginia | Florida | Iowa |
| --- | --- | --- | --- | --- |
| Mean | 0.0139 | 0.0082 | 0.0063 | 0.0105 |
| Standard Deviation | $\pm 0.0141$ | $\pm 0.0023$ | $\pm 0.0043$ | $\pm 0.0014$ |

**Figure S3:**
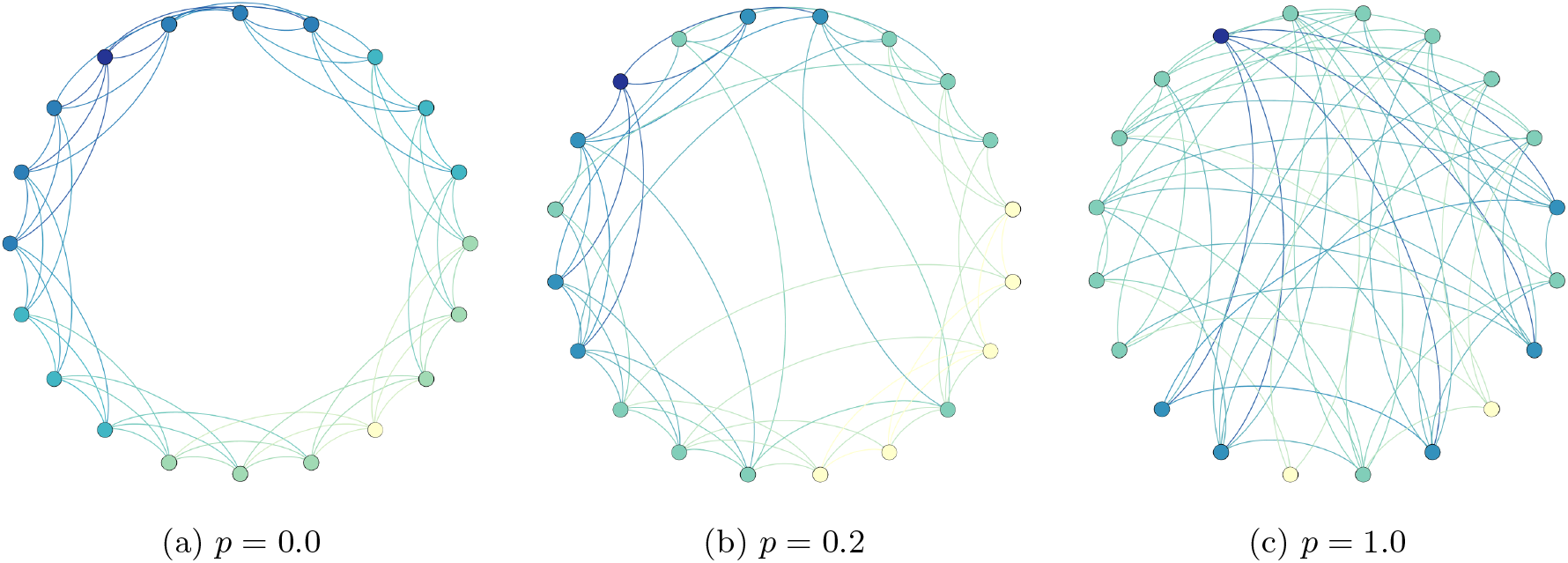
Representation of different network structures we obtain as we vary *p* the probability of rewiring edges. (a) Corresponds to the spatial network case, where only short edges are allowed; (b) represents the Facebook network structure, where short and long edges are exhibited; and (c) corresponds to a completely random scenario.

**Figure S4:**
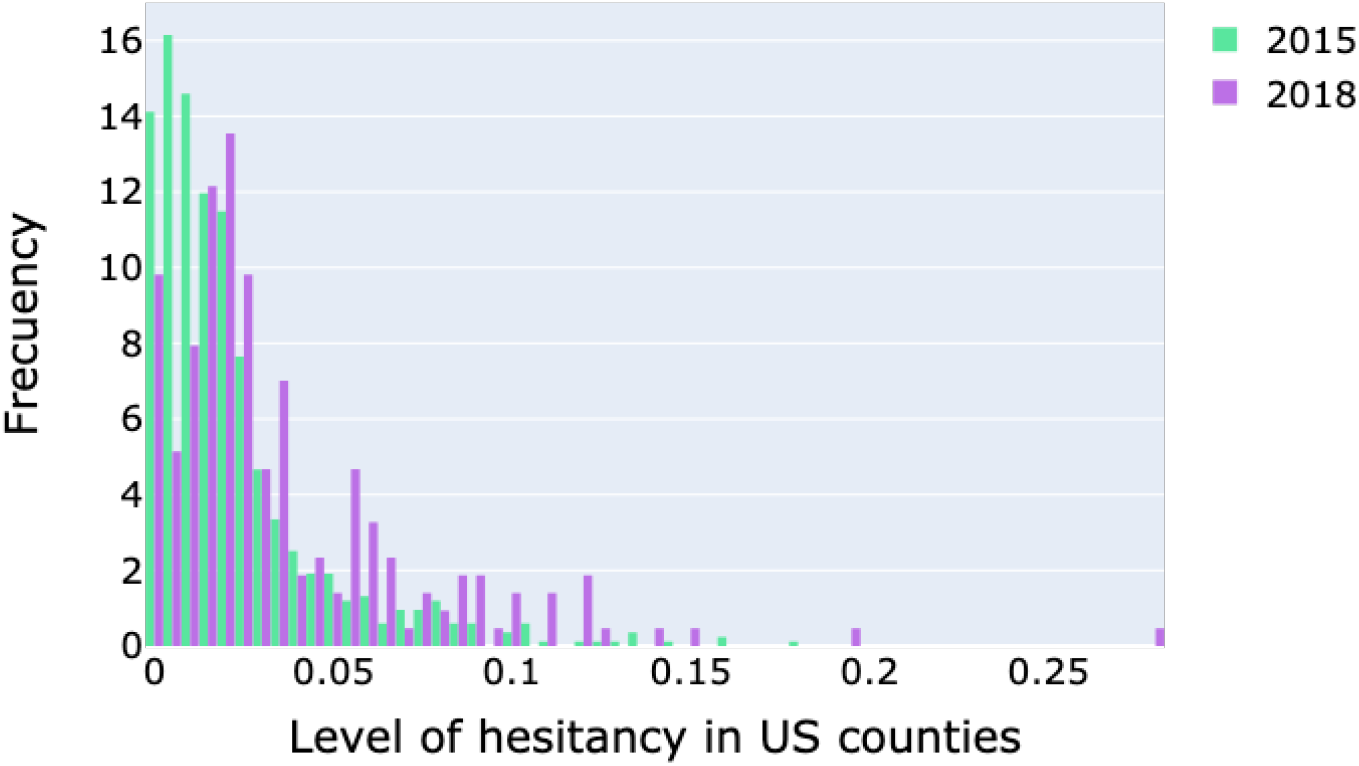
Distribution of vaccine hesitancy levels in the US for the years 2015 and 2018.

**Figure S5:**
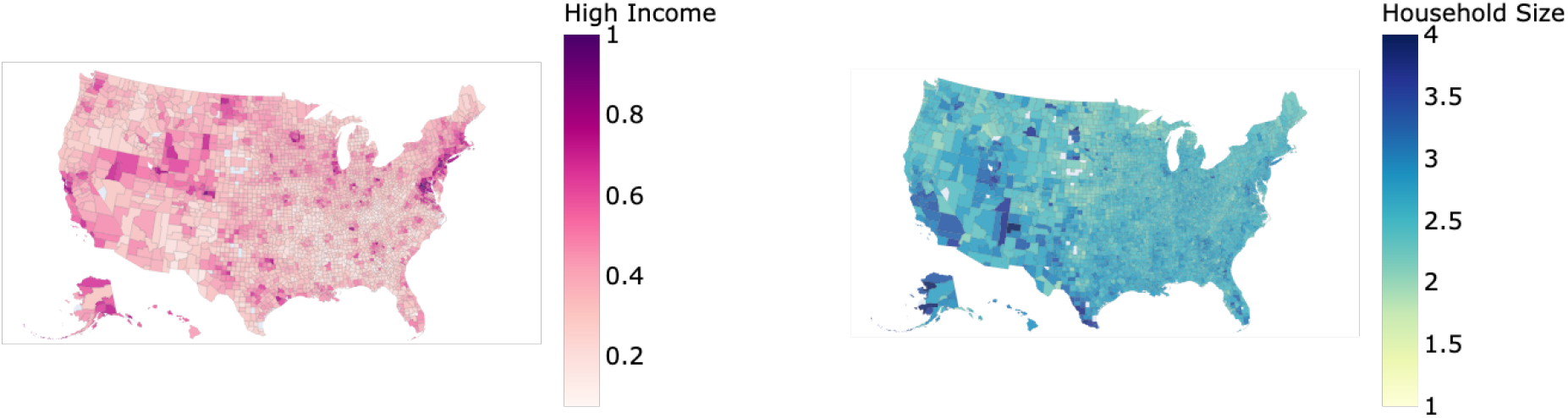
Distribution of traits associated with vaccination hesitancy in the united States for the year 2014.

**Figure S6:**
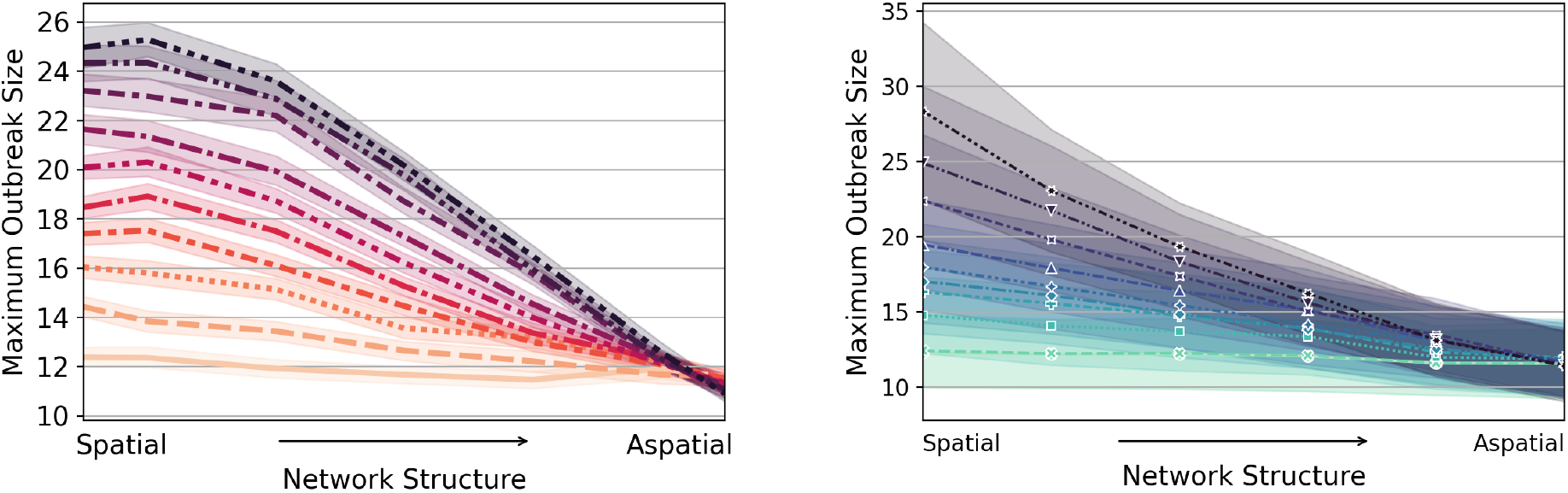
Heterogeneity in the levels of hesitant neighboring for the year 2015. By differentiating counties in vulnerable and protected states, we plot the level of hesitant each county is subjected.

**Figure S7:**
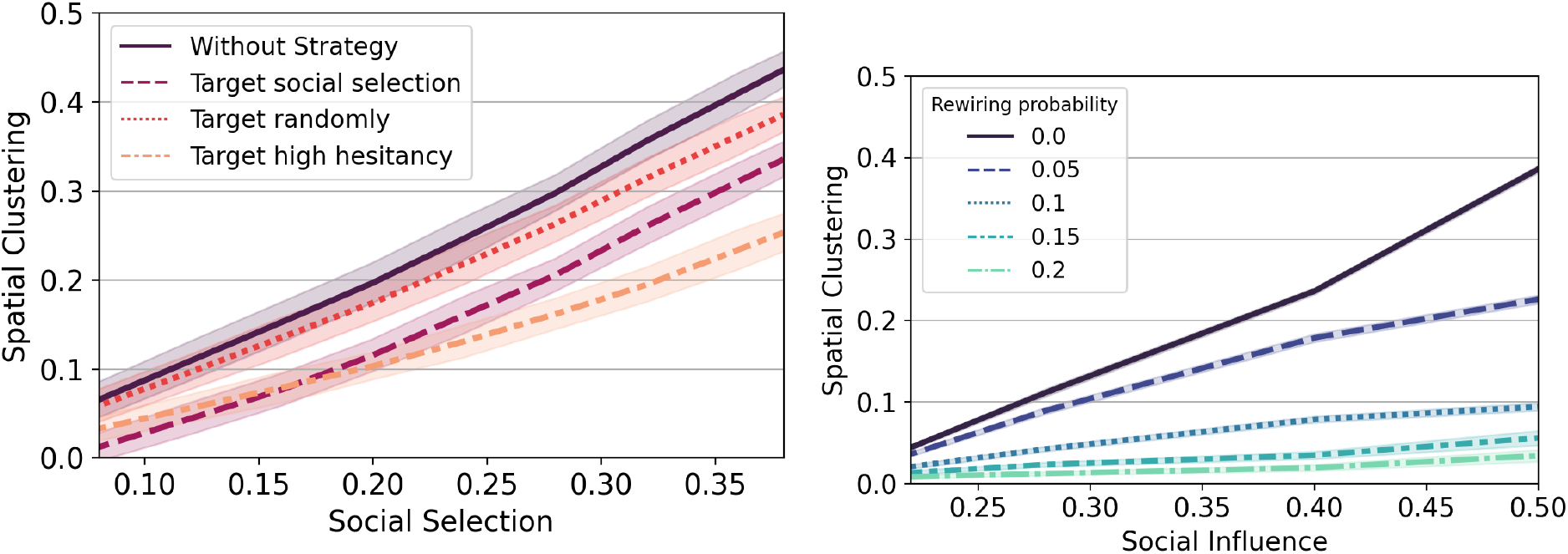
Theoretical results for the spatial clustering as a function of (a) social selection and (b) social influence, when mitigation strategies are applied. All parameters are set as in Fig.5

**Figure S8:**
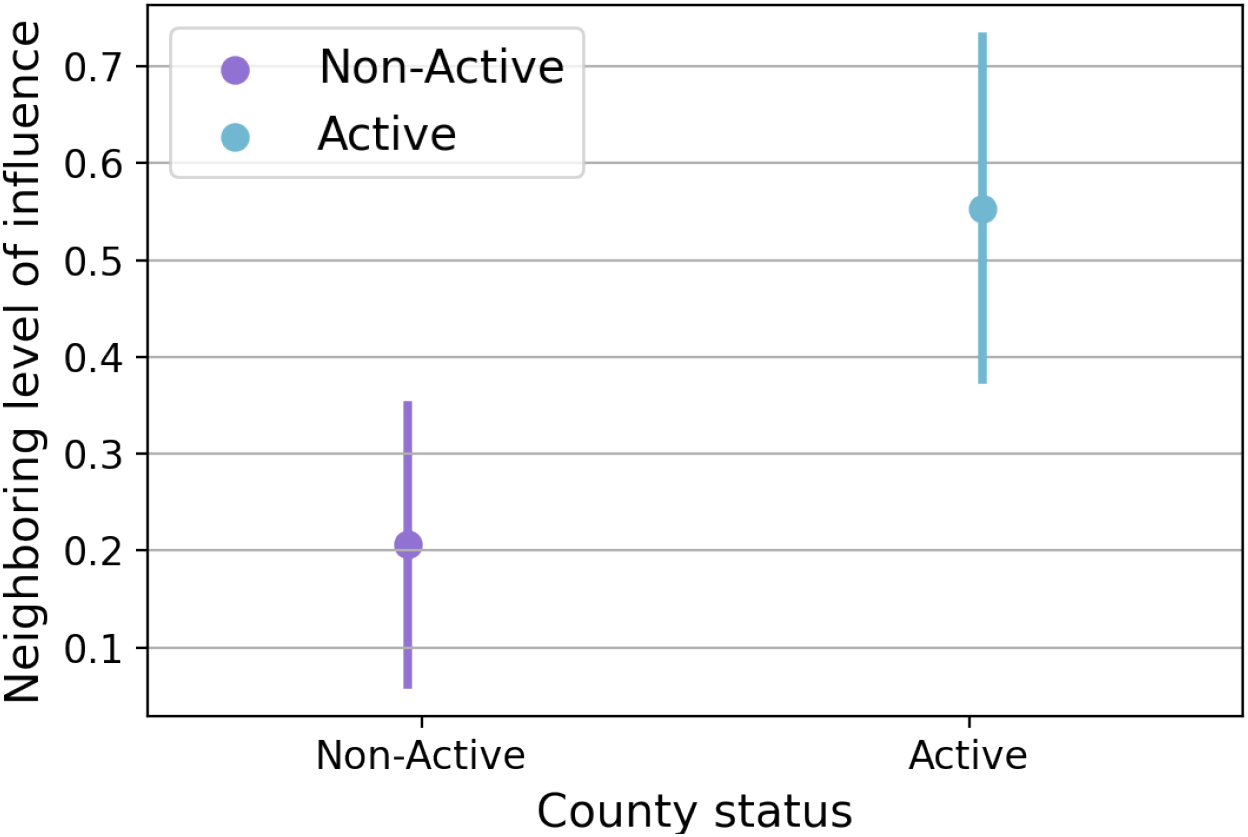
Heterogeneity in the levels of hesitant neighboring for the year 2015. By differentiating counties in vulnerable and protected states, we plot the level of hesitant each county is subjected.

**Figure S9:**
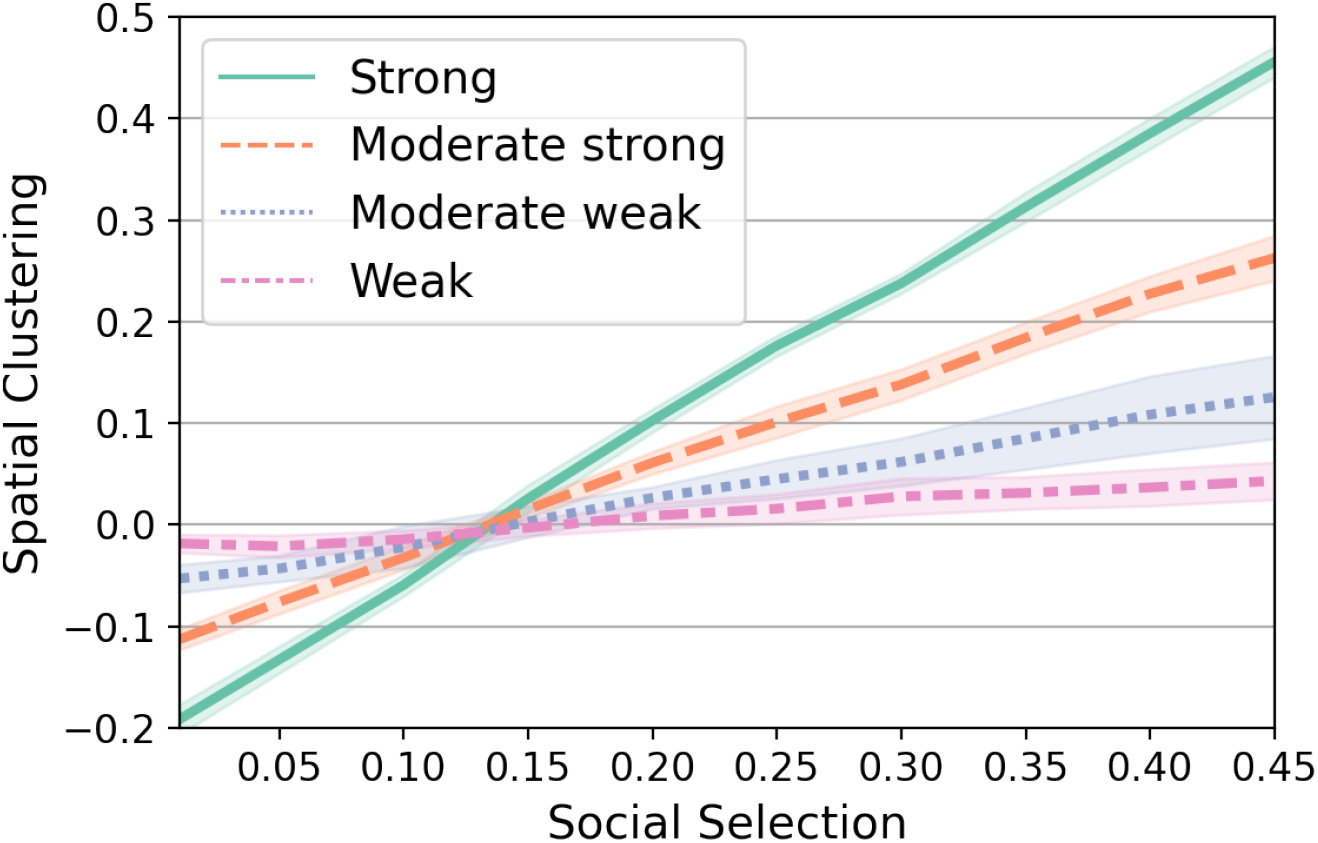
Correlation between traits and vaccine hesitancy as a function of social selection. As part of a sensitivity analysis, we explore how the spatial clustering is affected as the correlation between traits and hesitancy vary. To perform this different scenarios, we assign a vector of traits-hesitancy to each county, as explained in Methods section. We measure the correlation between each trait and hesitancy, considering that each county values is a point in the data. Then, we randomly chose two counties and swap their traits values in order to modify the correlation. And we proceed until we obtain the desire value.

